# A longitudinal seroconversion panel shows anti-SARS-CoV-2 antibody levels up to 6.5 months after vaccination with mRNA-1273 (Moderna)

**DOI:** 10.1101/2022.01.25.22269762

**Authors:** Francisco Belda, Oscar Mora, Rebecca Christie, Michael Crowley

## Abstract

**Background:** Vaccines have emerged as a crucial tool in combatting the COVID-19 pandemic particularly those based on SARS-CoV-2 S-protein mRNA. A crucial aspect of vaccine efficacy is the duration of antibody responses. In this study, a seroconversion panel was created to assess antibody responses to the mRNA-1273 vaccine over time (6.5 months).

**Methods:** Blood samples collected from 15 healthy adult participants prior to and up to 6.5 months after vaccination with the mRNA-1273 vaccine (Moderna). Serum from these blood samples were analyzed for anti-SARS-CoV-2 antibody activity by chemiluminescent immunoassay.

**Results:** The immunoassay results showed that one participant was positive for anti-SARS-CoV- 2 antibodies prior to vaccination indicating a prior infection. All participants showed a positive antibody response after the first vaccination. Highest antibody responses were seen after the second dose (41-45 days from the first dose). Subsequent samples collected at 69-75 days, 130-135 days and 221-229 days after the first vaccination showed positive responses but declining levels of anti-SARS-CoV-2 antibodies.

**Conclusions:** Declining antibody levels in these participants support the use of booster vaccination to increase antibody levels 4-6 months after the initial vaccine series and continued monitoring to assess the durability of COVID-19 vaccine responses. These results are in agreement with other studies showing antibody persistence but declining the antibody levels in the months after immunization with mRNA-based vaccines.

## Introduction

The SARS-CoV-2 pandemic has been a major public health challenge since its emergence in December 2019. [1] Efforts to contain COVID-19 have ranged from passive immunity in the form of convalescent plasma and monoclonal antibodies to pharmaceuticals to vaccines. Globally research has taken many approaches to vaccine development including mRNA, DNA, protein and non-replicating virus technologies. [2]

Two of the most advanced vaccines are those based on mRNA and nanotechnology, mRNA-1273 (Moderna) and BNT-162b2 (Cominarty, Pfizer-BioNTech). The mRNA-1273 vaccine is available for adults (≥18 years old) in the US under an Emergency Use Authorization (EUA) form the Food and Drug Administration (FDA). This vaccine is administered as two doses (100 µg) of pre-fusion stabilized spike protein mRNA given four weeks apart. [3] The BNT-162b2 vaccine has been approved by the FDA and is given as two doses administered 21 days apart. [4, 5] Both mRNA vaccines have been authorized by EUA for a third (booster) dose in people ≥ 18 years old, and people that are immunocompromised or at high risk of infection. [3, 4]

In this study, seroconversion panels were constructed from blood samples collected from study participants before and up to 6.5 months after vaccination and analyzed for anti-SARS-CoV-2 antibody responses. Seroconversion panels can have utility in the development of antibody assays and in the assessment of antibody responses over time. These assessments can include the duration of antibody responses and assessment of activity against emerging viral variants (e.g. B.1.1.529 [Omicron]). The goal of the study was to assess the anti-SARS-CoV-2 antibody response to mRNA-1273 vaccination over time.

## Material and Methods

### Seroconversion panel collection

The samples which make up these seroconversion panels were collected from volunteer healthcare workers at a Morristown, Tennessee (USA) hospital that had provided informed consent. This study was conducted under an approved IRB protocol ([SDP-003] Human Biological Specimen Collections: Diagnostic Investigational Review Board, Cummaquid, MA, USA). This study was conducted in compliance with all applicable regulatory guidelines.

This longitudinal panel was comprised of undiluted, unpreserved serum specimens collected from 15 participants between 25 February 2021 and 14 October 2021. The participants were 15 healthy adults 30-60 years of age who had received two injections of mRNA-1273 SARS-CoV-2 vaccine (100 µg) with an objective target of 28 days apart. There were nine male and six female participants in this group. All participants were white/Caucasian.

The samples for each participant were collected from before the first dose of the mRNA-1273 vaccine to 6.5 months after the second dose. In detail, samples are collected prior to the first vaccination (between 0 to 3 days: sample 1, pre-vaccination samples), prior to the second vaccination (between 2 to 7 days: sample 2), after the second vaccination (between 13 to 15 days; sample 3), 1.5 months after the second vaccination (between 41 to 45 days; sample 4), 3.5 months after the second vaccination (between 103 to 107 days; sample 5), and 6.5 months after the second vaccination (between 194 to 199 days; sample 6).

The serum samples were divided into 1 mL aliquots and frozen at −20°C until they were assayed. Samples were thawed at room temperature and mixed by inversion prior to testing. The serum samples were tested for anti-SARS-CoV-2 antibody activity with a chemiluminescent immunoassay (CLIA: Liaison SARS-CoV-2 IgG Assay, Diasorin, Inc, Saluggia, Italy: EUA approved).

## Results

Figure 1 shows the dynamic changes in antibody response against SARS-CoV-2 in a longitudinal seroconversion panel from 15 participants. Sample were collected before and after vaccination and up to 6.5 months after the second vaccine dose. In this figure, time 0 corresponds to the sample obtained before the first vaccination and the time shown for subsequent samples is number of days elapsed since collection of the pre-vaccinated sample.

**Figure 1.**
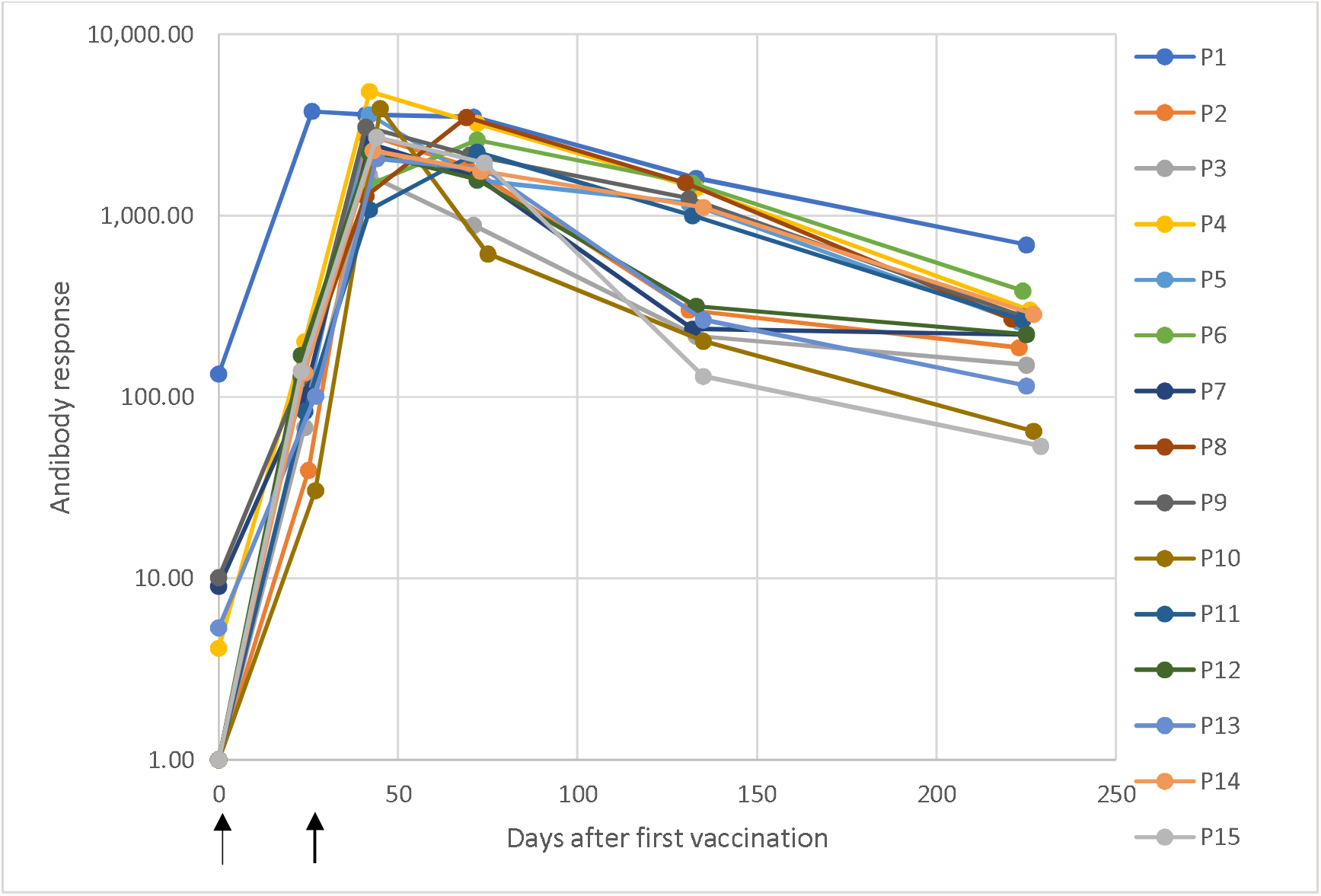
Dynamic changes in antibody response against SARS-CoV-2 in a longitudinal seroconversion panel from 15 participants (P1–P15) before and after vaccination with two doses of the mRNA-1273 SARS-CoV-2 vaccine (Moderna), indicated by arrows. Sample were collected prior to vaccination and up to 6.5 months from the second vaccine dose. These results were obtained using a chemiluminescent immunoassay and are expressed as arbitrary units/mL (AU/mL). Responses ≥ 15.0 units were considered positive. Time 0 correspond with the sample obtained before the first vaccination dose and the time of the subsequent samples are the elapsed days after the pre-vaccinated sample.

Testing of pre-vaccination samples gave the following results (Figure 1: day 0): 14 negative and one positive result (Participant 1). The presence of anti-SARS-CoV-2 IgG in this participant indicates that the participant was previously infected with the virus prior to collection of this sample.

Samples collected after the first injection showed 15 positive results (all participants). (Figure 1: 23-27 days). Samples collected after the 2nd dose (Figure 1: 41-45 days) also showed 15 positive results with the highest antibody responses. Samples collected 1.5 months post 2nd dose (Figure 1: 69-75 days) also showed 15 positive results. The antibody responses have started to decrease at this point. Samples collected 3.5 months post 2nd dose (Figure 1: 130- 135 days) showed positive results for all participants and a decrease in antibody responses compared with the previous sample. At 6.5 months post 2nd dose (Figure 1: 221-229 days) all participants showed positive results and the antibody response still decreasing compared with the previous sample collection.

## Conclusions

The results of this study showed that only one participant had an antibody response before the administration of the first dose of the mRNA-1273 SARS-CoV-2 vaccine. This positive response indicates a prior infection with SARS-CoV-2. All samples show positive results in antibody response after the first dose of the vaccine and all samples collected after 6.5 months post second dose were still positive. These results show a robust response to the first dose of the mRNA-1273 vaccine that is enhanced by the second dose of vaccine. The antibody response wanes over the post-vaccination period suggesting that humoral immunity declines in the months after vaccination.

These results agree with previous published observations which showed antibody persistence through 6 months after the second dose of the mRNA-1273 vaccine and assumes a steady decay rate over time. [6] A recent observational study looking at the real-world effectiveness of the mRNA-1273 vaccine (n=352,878 per group) showed that there was a small increase in SARS-CoV-2 infections in vaccinated individuals 4-5 months after vaccination. It is important to note that the infection rate in vaccinated participants remained well below the rate of infection in unvaccinated participants. Hospitalizations in the vaccinated group were less than 1/10 of those seen in the unvaccinated group and the number of deaths were far less in the vaccinated group (n=1) than in the unvaccinated group (n=25). [7]

## Data Availability

All data produced in the present work are contained in the manuscript.

## Acknowledgements

Michael K. James, Ph.D. is acknowledged for medical writing and Jordi Bozzo, Ph.D., CMPP for editorial assistance.

## Author Contributions

All authors have participated in the conception or design of the work, data collection and critical revision of the article.

